# Factors affecting trainees’ preferred timing of clinical clerkships during MD-PhD training

**DOI:** 10.64898/2026.05.10.26352845

**Authors:** Taylor P. Trentadue, Jessica Weng, Samantha M. Bouchal, Katherine E. Cornelius, Lisa M. Hurley, Leland S. Hu, Shannon P. Fortin Ensign, Rochelle R. Torgerson, Joseph J. Maleszewski, Bruce F. Horazdovsky, Scott H. Kaufmann, S. John Weroha, Lisa A. Schimmenti

**Affiliations:** Mayo Clinic Medical Scientist Training Program, Mayo Clinic, Rochester, Minnesota, United States; Mayo Clinic Graduate School of Biomedical Sciences, Mayo Clinic, Rochester, Minnesota, United States; Mayo Clinic Alix School of Medicine, Mayo Clinic, Rochester, Minnesota, United States; Mayo Clinic M.D.-Ph.D. Program, Mayo Clinic, Scottsdale, Arizona, United States

**Author notes:** co-first authors; contributed equally to this work (Taylor P. Trentadue, Jessica Weng). **Corresponding author** Lisa A. Schimmenti 200 First Street SW Rochester, Minnesota 55905.

**Keywords:** M.D.-Ph.D. program, medical education, physician-scientist training, undergraduate medical education

## Abstract

M.D.-Ph.D. programs in the United States have traditionally followed a “2-*n*-2” curricular model, with the graduate phase occurring between the pre-clerkship and clerkship portions of medical training. While well established, this format can limit trainee autonomy in shaping their physician-scientist training trajectories. In response, some programs have introduced a “3-*n*-1” model, allowing students to complete clerkships before beginning Ph.D. training. Our institution implemented multiple flexible curricula in 2017. Understanding why trainees choose one pathway is important as programs consider implementing more adaptable training structures. To investigate these factors, we surveyed M.D.-Ph.D. students at our institution, which offers multiple flexible curricular alternatives, about contributions to their curricular decisions. Responses supported that trainees weigh considerations across medical education, scientific development, and integrated physician-scientist training domains. Although the traditional pathway was a popular option, most students pursued one of the flexible pathways. Our findings suggest that introducing flexibility in the timing of undergraduate medical education and graduate training can support diverse educational, logistical, and personal priorities while maintaining the rigor of physician-scientist training. Offering multiple pathways empowers trainees to design trajectories that best fit their needs. Continued longitudinal studies are needed to assess the long-term impacts of flexible curricula on physician-scientist career outcomes.

## Introduction

Combined M.D.-Ph.D. programs have been training physician-investigators in the United States since 1964 (1). There are approximately 118 M.D.-Ph.D. programs in the United States and Puerto Rico, of which 53 are NIH T32 training grant-funded Medical Scientist Training Programs (MSTPs) (2, 3). Graduates from M.D.-Ph.D. programs comprise approximately 3.2% of in-training residents nationally (4). The traditional pathway for M.D.-Ph.D. dual degree trainees involves completing the pre-clerkship phase of medical training followed by three to five, hereafter *n*, years of Ph.D. training before returning to medical school for clerkships and post-clerkship experiences including sub-internships and clinical electives (5). In the traditional pathway, a prolonged interval between pre-clerkship and clerkship training can create barriers to physician-investigator training experiences (6, 7). Specifically, trainees returning to the clinic after *n* years of Ph.D. training may have or perceive deficits in their clinical knowledge or skills as well as less confidence in their clinical foundations (6, 8). From the scientific development perspective, trainees’ capacity to keep abreast of emerging literature or conduct laboratory work is limited during intensive clinical phases of medical school training. Trainees continually strive to balance the unique demands between their clinical and research development, and opportunities to strengthen this integrated skillset are important in early physician-scientist training.

There is growing interest in exploring curricular pathways tailored to the unique needs of trainees, including offering clerkships prior to or during Ph.D. training. Many M.D.-Ph.D. programs have trialed targeted programmatic offerings, including clinical re-immersion programs, to formalize the transition into clerkship training and address M.D.-Ph.D. trainee concerns (7, 9, 10). This curricular flexibility includes longitudinal clinical experiences in specialties related to laboratory work or career aspirations during the Ph.D. training years for specialty or subspecialty exploration, as well as incorporating Ph.D. rotations into pre-clerkship phase. However, the traditional “2-*n*-2” model—two years of pre-clerkship studies, *n* years of Ph.D. training, and two years of clerkship and post-clerkship studies—remains the predominant program structure (7, 11). As these flexible options are explored and implemented across programs, a deeper understanding of the factors that influence trainees’ decisions to pursue these different options is needed.

In 2017, the Mayo Clinic MSTP introduced flexible curricular pathways allowing trainees to engage with clinical clerkships and Ph.D. training on a timeline that can be individually tailored to a trainee’s interests and unique needs (**Figure 1**). These curricular pathways include the traditional “2-*n*-2” model, with all clerkships completed after Ph.D. training, and flexible models, which encompass all clerkships prior to Ph.D. training (“3-*n*-1”), a “split model” with any number of clerkships taken both prior to and after Ph.D. training, or any number of clerkships integrated prior to or during Ph.D. training.

**Figure 1.**
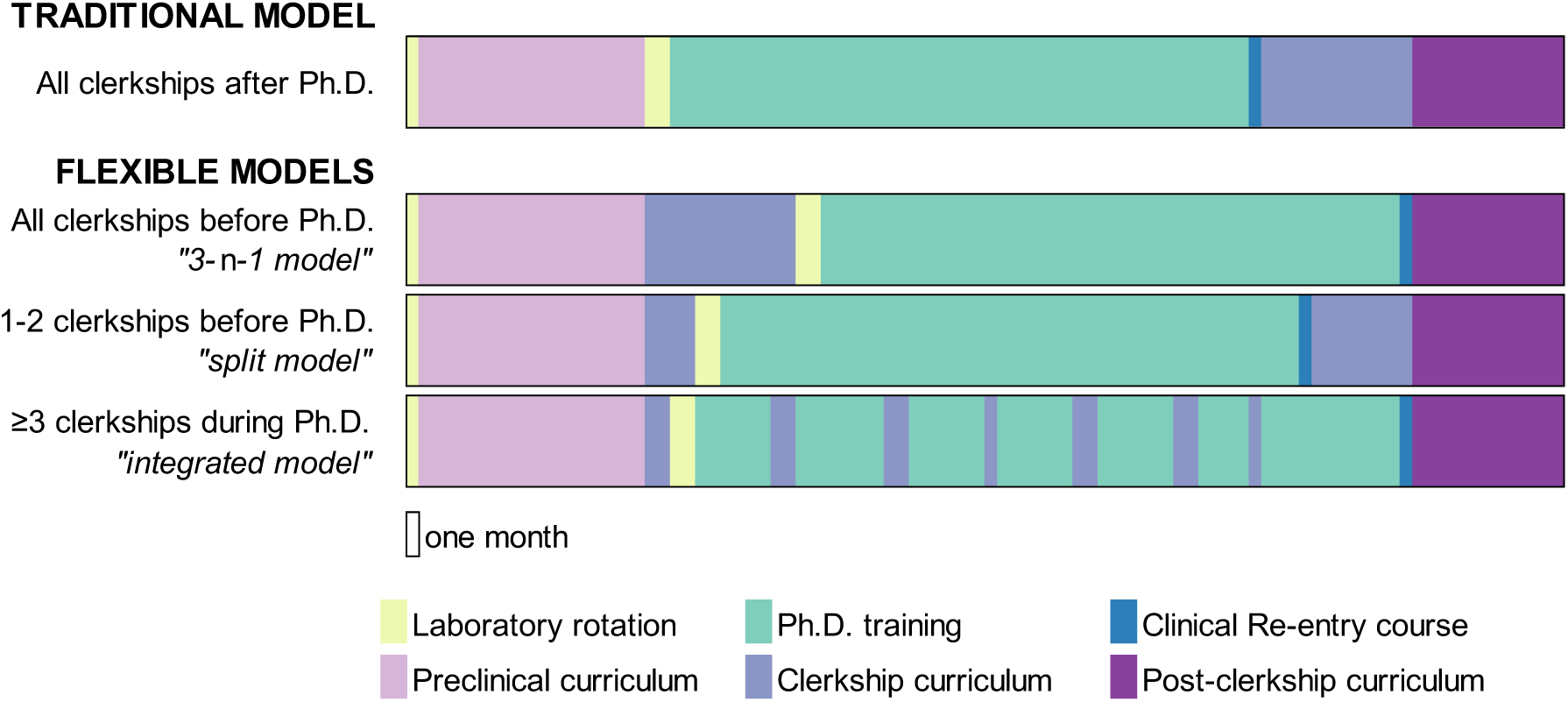
Curricular maps detailing the training pathways available to M.D.-Ph.D. trainees at our institution. The traditional model includes all clerkships after graduate training. The flexible curricular pathways include any number of clerkships before and during the graduate phase. In the integrated training model, the clerkship curriculum can be dispersed throughout the Ph.D. phase based on trainee preferences and clerkship availability per the medical school; one demonstrative example of a fully integrated training timeline is presented though distribution is customizable. The duration of the Clinical Re-entry course is one week at our institution.

The objectives of this study were to characterize factors influencing M.D.-Ph.D. trainees’ decisions to pursue curricular pathways offered at our institution. To accomplish this, we distributed a comprehensive questionnaire regarding curriculum pathway choice and factors influencing training pathway decisions to all M.D.-Ph.D. trainees at our institution. We hypothesized that logistical considerations from medical training, graduate training, and extracurricular or personal factors would inform trainees’ pathway decisions. Further, we hypothesized that the timing of clinical exposure during graduate training would inform and alter trainees’ clinical and research interests. Results of this analysis can inform the training strategies and curricular offerings at our institutional MSTP and provide insights to other M.D.-Ph.D. training programs.

## Results

### Stage of training and curricular pathway choice

An M.D.-Ph.D. trainee experience questionnaire was distributed to 63 trainees at our institution; all matriculated after the program introduced the flexible curricular pathway. For details regarding questionnaire content, please refer to Methods. Thirty-eight (60.3%) trainees participated. Respondents had completed median (25^th^ percentile, 75^th^ percentile) 3.5 (3.0, 6.0) years of training at the time of questionnaire completion; there was no significant difference in years completed between those who did versus did not participate (z=1.515, p=0.130; **Supplementary Figure 1**). Most respondents were enrolled in the graduate phase or completed their Ph.D. training at the time of questionnaire completion (n=23, 60.5%) (**Table 1**). There was no significant difference between years of M.D.-Ph.D. training completed in the traditional (3.0 [2.0, 5.3]) versus flexible (4.0 [2.8, 6.3]) curricular pathway groups (z=-0.653, p=0.514).

**Table 1.** Stage of training and curricular pathway choice. Data presented as n (%) or median (25^th^ percentile, 75^th^ percentile). n.s.: nonsignificant. † unpaired t-test. ‡ Fisher’s exact test or χ2 test. In the different pathway options, n refers to years of graduate training.

|  | Pathway<br>(n=38) |  | p value |  |
| --- | --- | --- | --- | --- |
|  | Traditional<br>(n=17) | Flexible<br>(n= 21) |  |  |
| **Years of M.D.-Ph.D. training completed** | 3.0 (2.0, 5.5) | 4.0 (2.5, 6.5) | n.s.† | 3.5 (2.0, 6.0) |
| **Phase(s) of M.D.-Ph.D. training completed or in progress \*** |  |  | n.s.‡ |  |
| Preclinical | 14 (82.3) | 15 (71.4) |  | 29 (76.3) |
| Some graduate school | 11 (64.7) | 12 (57.1) |  | 23 (60.5) |
| All graduate school | 1 (5.9) | 5 (23.8) |  | 6 (15.8) |
| Some clerkship phase | 1 (5.9) | 7 (33.3) |  | 8 (21.1) |
| All clerkship phase | 0 (0.0) | 7 (33.3) |  | 7 (18.4) |
| Some post-clerkship phase | 1 (5.9) | 4 (19.0) |  | 5 (13.2) |
| **Years of graduate school completed** | 1.0 (1.0, 3.0) | 1.0 (1.0, 2.8) | n.s.† | 1.0 (1.0, 3.0) |
| Missing § | 6 (35.3) | 9 (42.9) |  | 15 (39.5) |
| **Pathway** |  |  | <0.001‡ |  |
| Traditional model (2-n-2) | 17 (100.0) | - |  | 17 (44.7) |
| 1-2 clerkship(s) prior to graduate phase ("split model") | - | 8 (38.1) |  | 8 (21.1) |
| Preclinical + all clerkships prior to graduate phase ("3-n-1") | - | 7 (33.3) |  | 7 (18.4) |
| ≥3 clerkships during graduate phase ("integrated model") | - | 6 (28.6) |  | 6 (15.8) |
| **Clinical experiences during graduate phase** |  |  | n.s.‡ |  |
| Credited/graded \|\| | 3 (17.6) | 7 (33.3) |  | 10 (26.3) |
| Non-credited/ungraded # | 7 (41.2) | 7 (33.3) |  | 14 (36.8) |

\* As trainees are pursuing diverse curricular pathways, respondents indicated completed curricula or active enrollment. As some trainees have completed both some clerkships and some Ph.D. training, respondents were able to select multiple options and were counted more than once.
§ "Missing" indicates participants who omitted this question. Respondents had the option of choosing "Not applicable (N/A)" if they were enrolled in preclinical studies.
|| Credited options include clinical continuity electives during Ph.D. phase as well as the M.D.-Ph.D. Clinical Re-Entry course.

Among respondents, the traditional model was the most chosen singular pathway (n=17, 44.7%). The remainder of participants (n=21, 55.3%) completed one of the flexible pathways. Among the flexible pathway options, the split model of 1-2 clerkships prior to graduate training (n=8, 21.1%) and the 3-*n*-1 model of all clerkships prior to graduate training (n=7, 18.4%) were most frequently pursued; the remainder of trainees (n=6, 15.8%) chose to complete ≥3 clerkships during the graduate phase in the integrated training model (**Table 1**).

Almost half (n=18, 47.4%) of all respondents participated in clinical continuity experiences during graduate school; these included opportunities for credit (n=10, 26.3%) through the medical school and non-credited clinical experiences (n=14, 36.8%). There was no significant difference in the proportion of respondents participating in complementary clinical experiences during graduate training between trainees pursuing the traditional (n=7, 41.2%) versus flexible (n=11, 52.4%) pathways (χ^2^=0.473, p=0.492). All respondents (n=38, 100.0%) intend to pursue clinical residency after M.D.-Ph.D. training.

### Factors that influence pathway decision

Our primary analyses sought to understand factors that influence pathway decisions. We hypothesized that factors influencing decisions were within three broad domains: medical school, graduate school, and career development and personal factors. Additionally, we sought to understand whether curricular pathway decisions had a role in framing clinical and research interests.

When comparing respondents pursuing the traditional versus flexible curricular pathways, there were significant differences in clinical training-related considerations (p=0.047, **Figure 2A**) and the career development and personal factors domain (p=0.049, **Figure 2C**). We found no significant difference in graduate training-related considerations between respondents pursuing the traditional versus flexible curriculum (p=0.397, **Figure 2B**). Respondents pursuing the traditional pathway were more likely to consider the timing of fellowship grants (median agreement: 3 [2.5, 4] versus 2.5 [2, 4]) and publication logistics (median agreement: 4 [3, 4] versus 2 [2, 3]) as important factors.

**Figure 2.**
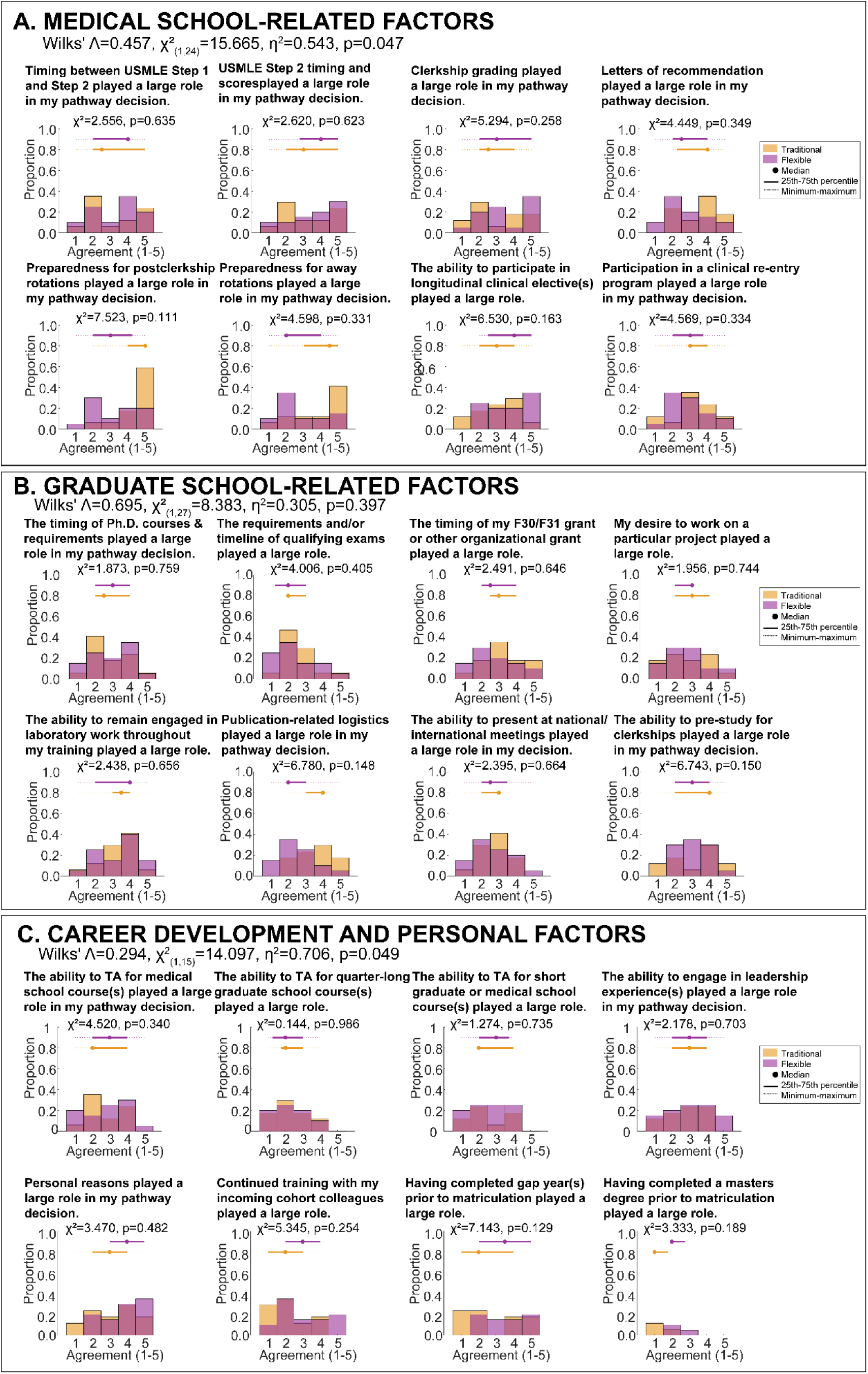
Proportion of respondents indicating levels of agreement with factors relating to curricular pathway decision-making stratified by curricular pathway option. (A) Factors related to medical school logistics. (B) Factors related to graduate school logistics. (C) Factors related to career development and personal logistics. Agreement was scored on a five-point Likert scale (1: strongly disagree, 2: disagree, 3: neutral, 4: agree, 5: strongly agree). The dots (solid line, dashed line) above each frequency plot represent the median (solid: 25^th^-75^th^ percentile, dashed: minimum-maximum) level of agreement. Section subtitles include statistics from one-way multiple ANOVA tests. Plot subtitles include the χ^2^ statistic comparing frequency of responses and associated p value for each individual factor.

The impact of pathway choice was not significantly associated with changes in clinical or research interests between respondents pursuing the traditional versus flexible pathways (p= 0.290, **Figure 3A**). Respondents pursuing the traditional pathway were more likely to endorse evolving clinical interests during the Ph.D. phase (p=0.045). In contrast, students in the flexible clerkship pathway were more heterogenous regarding clinical interest evolution, with trainees more frequently endorsing strong agreement or disagreement. Most respondents agreed that their pathway informed or refined their clinical interests in the traditional (median agreement: 4 [3, 4]) and flexible (median agreement: 4 [3, 5]) pathways.

**Figure 3.**
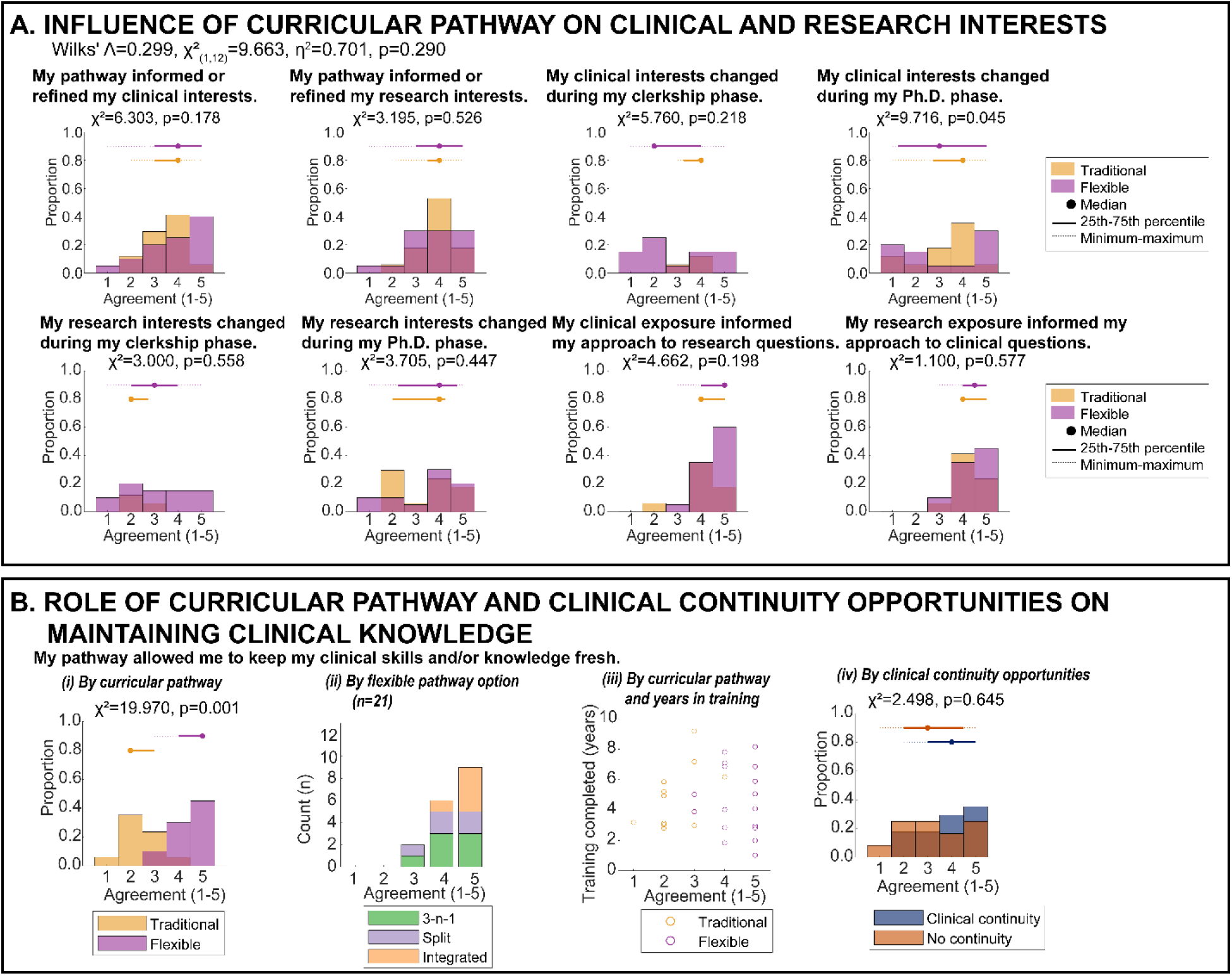
Levels of agreement with factors relating to clinical and research interests or knowledge stratified by curricular pathway. (A) Impacts of clinical and research training on career goals. (B) Maintenance and acquisition of clinical knowledge during training (i) by curricular pathway, (ii) by flexible pathway option, (iii) by years of training completed, and (iv) stratified by participation in longitudinal clinical opportunities. Agreement was scored on a five-point Likert scale (1: strongly disagree, 2: disagree, 3: neutral, 4: agree, 5: strongly agree). The dots (solid line, dashed line) above each frequency plot represent the median (solid: 25^th^-75^th^ percentile, dashed: minimum-maximum) level of agreement. Where statistics were performed, plot subtitles include the χ^2^ statistic comparing frequency of responses and associated p value.

Across all respondents, we sought to assess factors that were relatively more versus relatively less important in pathway choice. Within the medical school domain (**Figure 2A**), the individually perceived optimal timing of and performance on United States Medical Licensing Exam (USMLE) Step 2 (median: 4 [2, 5]) and preparedness for post-clerkship rotations and sub-internships (median: 4 [2.5, 5]) were important factors. In the graduate school domain (**Figure 2B**), the ability to remain engaged in laboratory work throughout all phases of M.D.-Ph.D. training was an important consideration (median: 4 [2.5, 4]), while timing of Ph.D. qualifying examinations was a less important (median: 2 [2, 3]). Although personal considerations were a key factor for most students (median: 4 [2, 5]), career development factors were less important, such as the ability to serve as a teaching assistant for full-term graduate (median: 2 [1.5, 3]), week-long intensive graduate (median: 2 [2, 4]) courses, or continue training with an incoming cohort of colleagues (median: 2 [2, 4]) (**Figure 2C**).

### Influence of pathways on training

The data underscore the dynamics between clinical and research experiences, suggesting that these complementary exposures influence interests across both domains. Regardless of curricular model, most respondents indicated that their chosen pathways influenced their clinical (median agreement: 4 [3, 4.75]) and research (median: 4 [2.5, 5]) interests (**Figure 3A**). This observation is consistent with the endorsed evolution of research interests during graduate training (median agreement: 4 [2, 4.5]). There was consistent agreement among M.D.-Ph.D. trainee respondents that their clinical exposure informed their approaches to research questions (median agreement: 4.5 [4, 5]) and that their research exposure informed their approaches to clinical questions (median agreement: 4 [4, 5]).

There was a significant difference in perceived maintenance of clinical knowledge between curricular pathway chosen (**Figure 3B**). Respondents who pursued a flexible pathway more strongly agreed that their pathway allowed them to keep their clinical skills and knowledge current (median agreement: 5 [4, 5]) compared to trainees pursuing the traditional pathway (median agreement: 2 [2, 3]) (p=0.001). There was no significant difference in perceived clinical performance between those pursuing or deferring clinical continuity experiences (p=0.645) (**Figure 3B**).

### Preliminary insights into each flexible pathway option

The flexible pathway options were chosen by 21 (55.3%) respondents, with eight (38.1%) selecting the split model, seven (33.3%) the 3-*n*-1 model, and six (28.6%) the integrated model. Given the small number of trainees in each pathway, formal statistical analyses were not performed. The frequencies of agreement across the flexible pathway options within the medical school (**Figure 4A**), graduate school (**Figure 4B**), and personal factors (**Figure 4C**) domains provide preliminary insights into considerations for each unique option. Similarly, the frequencies of agreement for the impact on flexible pathways on training trajectories (**Figure 4D**) offer preliminary information regarding the role of flexible pathways in shaping physician-scientist development. All participants pursuing the integrated flexible model agreed or strongly agreed (median agreement: 5 [4.75, 5]) that the pathway allowed for maintenance of clinical skill and knowledge (**Figure 3B**).

**Figure 4.**
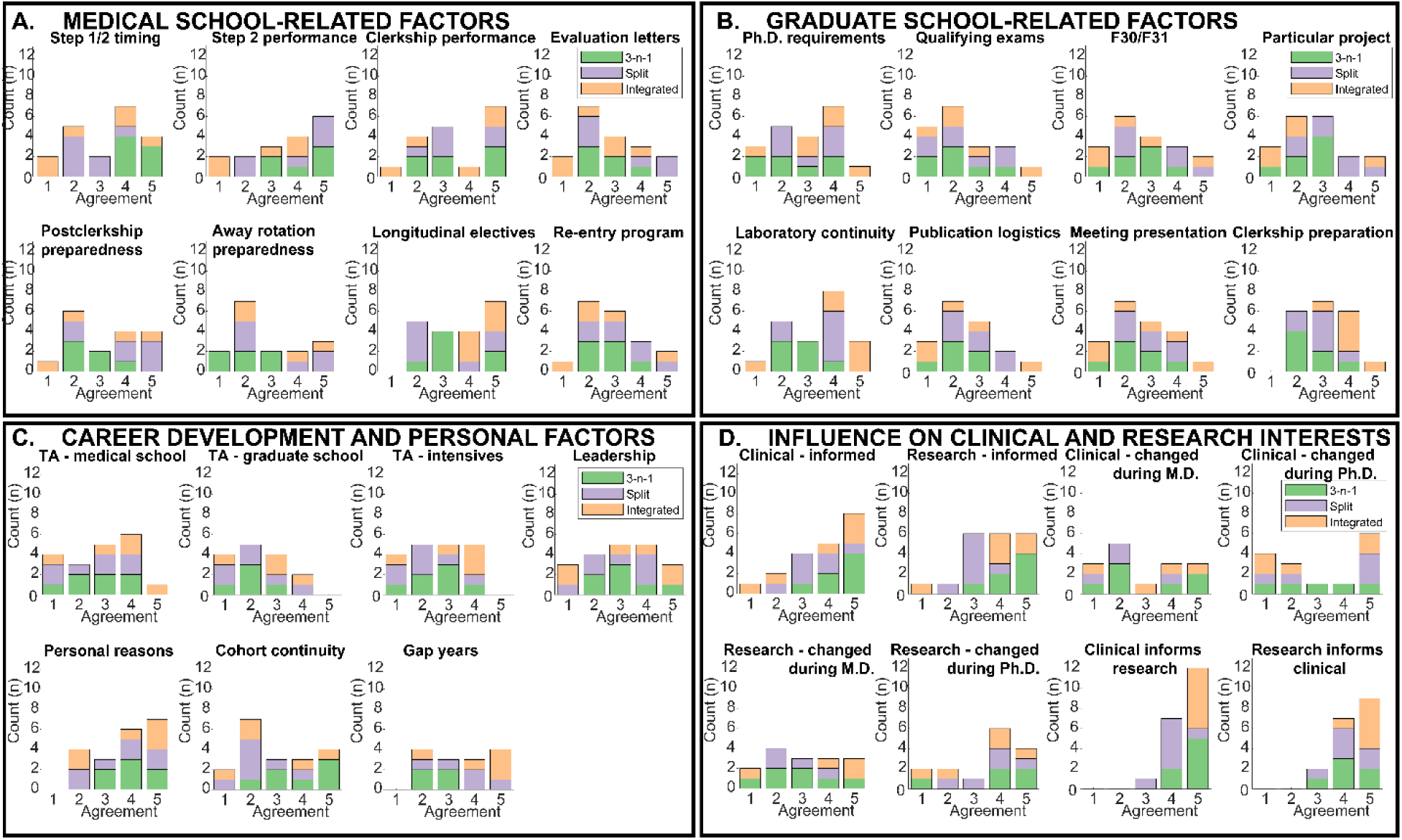
Levels of agreement with factors relating to (A) medical school, (B) graduate school, and (C) personal factor domains as well as (D) role of pathway on influencing physician-scientist development among the 21 (55.3%) participants who selected a flexible pathway option. Given the small sample sizes, these data are descriptive and exploratory. The short-hand titles used in each subplot correspond to the prompts as presented in **Figures 2A – 2C** and **Figure 3A**.

## Discussion

The structures of M.D.-Ph.D. programs, which conventionally follow a traditional “2-*n*-2” format, pose challenges for trainees re-integrating into medical training and striving to balance the demands of both physician and scientist developmental trajectories. Leaders at dual-degree training programs are implementing innovative approaches to address these needs. According to a survey of 92 M.D.-Ph.D. Programs in the United States, 65.2%, 13.0%, and 21.7% of curricula follow the traditional 2-*n-*2 pathway, all clerkships prior to Ph.D. training in the 3-*n*-1 model, and the split model of some clerkships prior to graduate training, respectively (12). However, few programs offer trainees the individually-tailored opportunity to select among these options or to integrate clerkships throughout Ph.D. training. Thus, the current study sought to understand the factors that influenced pursuit of the traditional versus a flexible training curriculum when multiple offerings are available.

As there is no standardized curriculum for M.D.-Ph.D. programs across the country (12), curricular flexibility in M.D.-Ph.D. programs allows trainees to tailor their academic trajectories to their unique interests and needs while maintaining rigor across medical and scientific development. This information may be useful to M.D.-Ph.D. program leadership who may be considering implementation of similar curricular flexibility at their institutions prospective as well as M.D.-Ph.D. applicants hoping to explore the nuances of different training pathways. Further, for a prospective or early-stage trainee, awareness of factors that may influence curricular pathways may be helpful in making their own training decisions.

At our institution’s M.D.-Ph.D. program, approximately half of trainees (44.7%) pursued the traditional model, with the remainder (55.3%) selecting a flexible curricular option. Of the flexible pathways, the split model of one to two clerkships prior to graduate training or the 3-*n*-1 model of all clerkships prior to graduate training were most often chosen by trainees. By offering flexible physician-scientist training options—namely, the traditional model as well as the option to complete clinical clerkships before or during the graduate phase—programs can support a variety of goals and allow trainees to individualize their educational paths.

We sought to understand factors that may influence trainees to choose curricular pathways. Between traditional and flexible pathways, significant differences were observed within clinical training, career development, and personal factor domains. Within the medical school consideration domain, USMLE Step 2 scores and preparedness for post-clerkship clinical experiences were considered important to strongly important factors across trainees in both pathways. This may be especially important considering the recent transition of USMLE Step 1 scoring from numeric to pass-fail. Specifically, the scoring change may relate to increased emphasis on other portions of trainees’ residency applications; these include USMLE Step 2 scores, meaningful research experiences, clerkship grades, medical school reputation, and letters of reference (13–17).

Among the career development and personal factors domain, personal factors were considered a relatively more important factor. This may suggest that trainees appreciate the option of a flexible curricular model to allow them to integrate other life domains into training. Future qualitative studies may consider querying themes within this broad domain to better support physician-scientist trainees. The ability to serve as a teaching assistant for medical school or graduate schools and pursue continued training with incoming cohort colleagues were considered less important influences on pathway selection. This may be because teaching opportunities can be pursued with relative flexibility at our institution for trainees interested in pursuing medical education leadership skills. While there was not a significant difference in the influences of graduate training-related domain considerations between curricular pathways, the ability to engage in laboratory work throughout training was identified as more important factor for pathway decision-making in all respondents; this highlights that all trainees are keen to find timelines that encourage preparedness for an integrated physician-scientist career.

All respondents reported that clinical exposure strongly informed their approaches to research questions and that research experiences informed their approaches to clinical questions, independent of curricular pathway chosen. This supports the concept that M.D. and Ph.D. training experiences are synergized, with benefits regardless of the curricular model. We found a significant difference in evolving clinical interests between those pursuing the traditional versus flexible pathways. Those in the flexible pathway were more likely to strongly agree or strongly disagree that their clinical interests changed during graduate training, whereas trainees in the traditional model were more likely to agree that their clinical interests shifted while research interests remained more consistent. This may indicate that trainees with very clear training goals or those exploring multiple specialties perceive benefits from flexible training timelines, whereas those who chose the traditional pathway may be more confident about preferred research areas.

Many respondents pursued a flexible pathway option, allowing them to complete at least one clerkship experience prior to or during the Ph.D. phase. The opportunity to pursue at least one clerkship allows trainees to gain insight into their roles and expectations during clinical training, which may alleviate concerns about transitioning between phases or allow them to more robustly participate in longitudinal clinical experiences. Compared to their categorical M.D. colleagues, M.D.-Ph.D. trainees may experience or perceive deficits in clinical skills or performance in clinical environments after multiple years in the laboratory. In the current study, trainees pursuing the flexible curricular model were more likely to agree that their pathway would allow them to keep their clinical skills or knowledge more current relative to trainees pursuing the traditional model.

All respondents to this questionnaire participated in complementary clinical experiences during graduate school training, suggesting that these opportunities are valuable to physician-scientist trainees. Clinical continuity experiences represent an important means by which M.D.-Ph.D. trainees can maintain clinical skills throughout their tenure in the program. While all respondents pursued elective clinical continuity opportunities during graduate training, our results may suggest that flexible pathways encourage more sustained or perceived clinical proficiency. Since trainees in the traditional pathway may perceive reduced retention of clinical knowledge and skills (6, 8), longitudinal follow-up questionnaires at the conclusion of M.D-Ph.D. training will be valuable. At our institution, the first M.D.-Ph.D. cohort with flexible options graduated in May 2025, limiting our ability to query perceptions at the time of transition to residency as well as performance in the National Resident Matching Program.

### Perspectives from M.D.-Ph.D. and medical school leadership

From a leadership standpoint, offering multiple pathways allows trainees to be fully engaged in decision-making related to their educational pathways. Whether a trainee chooses a traditional “2-*n*-2” pathway or one of the flexible pathway options, they are deciding and, in the “act of choosing,” trainees become active participants in their personal curricular development. We propose that this “act of choosing” gives trainees ownership of their career development and academic choices. From an administrative standpoint, a program with trainees following different curricula provides several logistical challenges. These are mitigated by the development of milestone tracking software to ensure that trainees are meeting appropriate milestones across both the medical and graduate phases of training regardless of the pathways they pursue.

Clinical experiences and “re-entry” programs for M.D.-Ph.D. trainees have been designed to maintain clinical skills during graduate training (9); these have the potential to improve clerkship performance (10). At our institution, a 40-hour practical refresher course is offered to M.D.-Ph.D. trainees intending to transition back to medical school during or immediately after graduate school. Similarly, longitudinal clinical engagement courses or experiences can be integrated during the Ph.D. phase. To promote clinical experiences during the Ph.D. phase, our institution’s M.D.-Ph.D. program offers three opportunities for clinical continuity as well as a structured clinical re-entry program intended to be taken at the end of the Ph.D. phase. It may be useful to formally acknowledge these clinical experiences on transcripts to demonstrate sustained clinical exposure during graduate training (7).

Since the flexible pathway options were implemented at our institution in 2017, the structure of pre-clerkship training nationally continues to evolve. Most medical schools now offer a 12-to-18 month pre-clerkship phase, a modification from the traditional two-year pre-clerkship structure. Our institution transitioned from a two-year to an 18-month pre-clerkship curriculum beginning in the 2021-2022 academic year, meaning that half of the cohorts included in this study matriculated under each pre-clerkship training paradigm. This modification to the “2-*n*-2" paradigm may have additional influence on flexible pathway selection, particularly as students may wish to align the start of graduate school with the new academic year.

### Strengths

This study evaluates curricular choices in a novel M.D.-Ph.D. curricular choice model, with options for students to complete clerkships prior to or during the graduate phase as well as longitudinal clinical experiences taken for medical school clinical elective credit. Although respondents may have completed part of their medical or graduate training at any of the three Mayo Clinic sites located in Minnesota, Arizona, or Florida, all students followed a unified curriculum established by the medical and graduate schools, which is consistently implemented across all locations. Additionally, oversight of the M.D.-Ph.D. program is coordinated centrally, ensuring consistency in training and administration throughout all sites.

### Limitations

The findings of the current study must be viewed in the context of its limitations. The flexible curricular pathway was implemented for the incoming cohort of 2017; as such, all respondents were actively enrolled in M.D.-Ph.D. training at the time of this study. Long-term analyses—such as differences in time to degree, residency placement, or other career outcomes—as well as medical school performance, including clerkship performance or USMLE scores, could not be performed in a statistically meaningful manner at this time. Given the unique curricular model, trainees at only a single institution were queried. Finally, the response rate (60.3%), while robust for an e-mail-based notification system with a restricted number of reminders based on institutional review board protocols, may not be fully reflective of the entire trainee population and the breadth of factors influencing pathway decisions.

## Conclusion

The decisions involved in curricular pathways are diverse and multifactorial. They reflect a variety of concerns in medical, scientific, and synergized physician-scientist program training requirements, as well as plans for future training and career trajectories. Overall, most trainees pursued some variant of a flexible curriculum. Emphasis should be placed on curricular pathway structures that train excellent physician-scientists and support diverse physician-scientist trajectories (5, 18). At our institution, trainee experiences suggest that offering multiple curricular pathway options allows learners to tailor curricula to their unique needs.

## Methods

### Study approval

This study involved sending a questionnaire about factors influencing curricular pathway decisions to all M.D.-Ph.D. trainees at our institution. The custom questionnaire was sent via e-mail to all M.D.-Ph.D. trainees at the Mayo Clinic – Rochester, Minnesota campus following an IRB-approved protocol. The questionnaire was administered via Research Electronic Data Capture (REDCap), an institutionally-supported HIPAA- and FERPA-compliant data collection and management software (21). Written informed consent was obtained from participants prior to participation. Respondents were remunerated with a meal voucher of nominal value to the hospital’s cafeteria.

Following closure of the REDCap response window, an anonymized report containing binary response status and year of matriculation was generated. This was used to compare differences in stage of training, as estimated by years since matriculation, between those who did and did not participate in the study.

### Questionnaire design

The questionnaire (**Supplementary Appendix 1**) queried stage of training and training completed to date, participation in longitudinal clinical experiences, and sources of advice on model selection. It captured trainees’ curricular models, influence of complementary training on clinical and research domains, influence of logistical concerns on pathway, and influence of specialty-specific concerns on pathway. Questions pertaining to factors that may or may not influence curricular decisions and residency competitiveness were by a five-point Likert scale (1: strongly disagree, 2: disagree, 3: neutral, 4: agree, and 5: strongly agree). Participants had the option of selecting “not applicable (N/A)” if the factor did not apply to them; these responses were excluded from analysis.

### Primary analysis

Our primary hypothesis was that factors influencing selection of traditional versus flexible curricular pathway would differ between those in each group. These factors included considerations in the medical school domain, the graduate school domain, and career development and personal domain:

- The **medical school domain** included factors such as timing between USMLE Step 1 and Step 2, performance on and timing of USMLE Step 2, clerkship grading, letters of recommendation, preparedness for post-clerkship rotations, preparedness for away rotations, ability to participate in longitudinal clinical electives, and ability to participate in a clinical re-entry program.
- The **graduate school domain** included factors such as timing of required Ph.D. coursework and requirements, requirements and/or timeline of qualifying exams, timing of F30/F31 grant or other organizational/institutional grants, desire to work on a particular project, ability to remain engaged in laboratory work throughout training, logistics related to publications, ability to present at conferences or meetings, and ability to pre-study for clerkships.
- The **career development and personal factor domain** included factors such as ability to be a teaching assistant for medical school and graduate school courses, ability to participate in leadership experiences, personal reasons, continued training with incoming cohort colleagues, having completed gap year(s) prior to matriculation, and having completed a master’s degree prior to matriculation.

Additionally, we hypothesized that the curricular pathway would have different effects on informing clinical and research interests. Results are shown by median (25^th^ percentile, 75^th^ percentile) to estimate the importance of these factors to curricular decision-making.

### Secondary analysis

We hypothesized that certain factors would be of greater importance than others to all trainees, independent of curricular pathway. We calculated the medians of ordinal, Likert scale data across respondents to assess the relative importance of each factor (19, 20). We categorized relatively more and relatively less important factors as those with median scores ≥4 or ≤2, representing medians within the agreement to strong agreement range versus disagreement to strong disagreement range, respectively.

### Statistical analysis

Categorical data were analyzed by frequencies and percentages. Summary statistics (median [25^th^ percentile, 75^th^ percentile]) were reported for ordinal data (19, 20). Unpaired t-tests or Wilcoxon rank sum tests were used for continuous variables; χ^2^ or Fisher’s exact tests were used for categorical variables. To analyze the effect of each consideration domain—medical school logistics, graduate school logistics, and career development logistics—on pathway decisions and the influences of pathway decisions on clinical and research training trajectories, ordinal data were treated as continuous variables, and one-way multiple ANOVA tests were performed. To complement this analysis, all ordinal question-level data were analyzed using unpaired Wilcoxon rank sum tests and reported in **Supplementary Appendix 2**. As this study is exploratory in nature, no adjustments for multiple comparisons were applied.

Given the small sample size of trainees within each of the flexible curricular pathways, formal statistical analyses were not performed. Instead, frequencies of agreement are presented as descriptive results to offer preliminary insights into factors relating to selection of a particular flexible trajectory. As no statistical tests were performed, significance is not reported.

## Data availability

Individualized data are not available to protect anonymity. Aggregate data may be available from the authors upon reasonable request.

## Author contributions

T.P.T.: conceptualization, methodology, investigation, formal analysis, visualization, writing – original draft, writing – review & editing. J.W.: conceptualization, methodology, investigation, formal analysis, writing – original draft, writing – review & editing. S.M.B.: investigation, writing – original draft, writing – review & editing. K.E.C.: data curation, writing – review & editing. L.M.H.: project administration, resources. L.S.H.: writing – review & editing. S.P.F.E.: writing – review & editing. R.R.T: writing – review & editing. J.J.M.: writing – review & editing. B.F.H.: writing – review & editing. S.H.K.: funding acquisition, writing – review & editing. S.J.W.: supervision, project administration, writing – review & editing. L.A.S.: conceptualization, methodology, investigation, funding acquisition, supervision, project administration, writing – review & editing.

T.P.T. and J.W. are co-first authors and listed alphabetically by last name.

## Acknowledgements

The authors thank Erin Triplet, M.D., Ph.D., for initiating the curricular transitions guide during her tenure as a Medical Scientist Training Program (MSTP) trainee. The authors thank the current trainees who have provided early input on curricular pathway decisions in both written form and panel discussions since the introduction of the flexible curricular pathway in 2017.

