## Supplementary Appendix 1 for "Factors affecting trainees’ preferred timing of clinical clerkships during MD-PhD training"

### Factors affecting M.D.-Ph.D. curriculum model selection

Please complete the survey below.

You are being asked to participate in a research study called "Factors Affecting M.D.-Ph.D. Curriculum Model Selection: A Single Institution's Experience". The purpose of this research is to characterize factors influencing trainees' decisions to pursue different M.D.-Ph.D. curricular pathways. You have been asked to take part in this research because you have been identified as a student in the Mayo Clinic M.D.-PhD. Program.

Study participation involves completing an electronic questionnaire that does not include any direct identifying information. Although the results of this study may be published, no information that could identify you or protected health information will be collected as part of this study.

Participating in research is completely voluntarily. Please understand that if you choose not to participate there will be no impact on your standing in the educational program, evaluation, or future employment opportunities at Mayo Clinic or your relations with the Mayo Clinic Medical Scientist Training Program, Mayo Clinic Graduate School of Biomedical Sciences, or Mayo Clinic Alix School of Medicine.

To maintain your anonymity, no data will be accessed until after the recruitment period of one month.

What is your gender identity?	1. Female

**DEMOGRAPHICS**

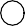

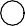

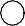

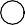

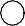

2. Male
3. Nonbinary / gender non-conforming
4. Other
5. Prefer not to answer

What is your current age?	1. 20-24 years

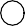

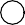

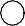

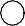

2. 25-29 years
3. 30-34 years
4. 35+ years

What is your racial / ethnic identity? Please choose
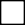
 1. American Indian or Alaska Native all that apply.
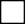
 2. Asian

3. Black or African American
4. Native Hawaiian or Other Pacific Islander
5. White
6. Hispanic
7. Middle Eastern or North African
8. Other
9. Prefer not to answer

1. Minnesota - Rochester	2. Arizona	3. Florida

**For each stage of your training, which campus were you on or which campus do you plan to be**

**on?**

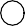

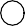

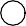

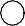

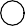

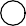

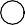

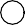

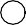

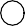

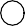

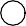
Preclinical Graduate Clerkships Post-clerkships

For the question below, please refer to the following map.

What area of the United States are you from?
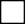
 1. New England

2. Middle Atlantic
3. South Atlantic
4. East North Central
5. East South Central
6. West North Central
7. West South Central
8. Mountain
9. Pacific
10. International

Do you have dependents?	1. Yes

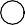

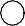

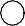

2. No
3. Prefer not to answer

How many years of M.D.-Ph.D. training have you completed?

**STAGE OF TRAINING**

(Include current year. Whole numbers only.)

What phase(s) of M.D.-Ph.D. have you completed or are
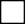
 1. Preclinical

you actively enrolled in?
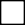
 2. Some graduate school

3. All graduate school
4. Some clerkship phase
5. All clerkship phase
6. Some post-clerkship phase

How many years of graduate school have you completed?

(Include current year. Whole numbers only.)

What is/was the duration of your preclinical	1. 18 months (incoming class of 2021+)

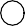

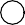

curriculum?	2. 24 months (incoming classes until 2020)

Which general pathway did you pursue / will you
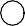
 1. Traditional model (preclinical, all or most of pursue?	Ph.D., clerkship + post-clerkship)

2.
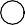
Traditional model + 1-2 clerkship(s) prior to Ph.D.
3.
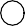
Traditional model + 1-2 clerkship(s) prior to formal clerkship transition
4. Preclinical + all clerkships prior to Ph.D.

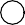

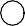

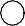

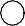

5. Hybrid model (3-5 clerkships during Ph.D.)
6. Hybrid model (6-7 clerkships during Ph.D.)
7. Hybrid model (all clerkships during Ph.D.)

Please select your Ph.D. track.	1. BMB

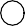

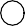

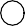

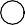

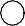

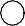

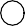

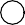

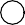

2. BMEP
3. CTS
4. IMM
5. MPET
6. NSC
7. REGS
8. VGT
9. Undecided
1. Yes	2. No

**Which clerkship(s) did you complete before beginning your Ph.D. phase OR during your Ph.D.**

**phase?**

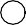

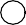
N/A: I am in preclinical training and do not have clerkship enrollment information yet.

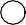

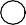

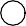

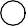

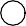

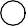

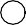

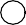

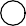

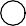
Emergency Medicine Family Medicine Internal Medicine Neurology

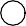

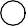

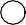

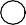
Obstetrics and Gynecology Pediatrics

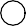

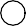
Psychiatry Surgery

Have you completed any of the following clinical
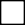
 1. MDPH 6100

experience(s) since starting your Ph.D. phase?
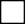
 2. CLNX 7640

3. Clinical re-entry with Dr. Weroha
4. Non-credited experiences (e.g. volunteer experiences, shadowing, global health, etc.)

How many times have you completed MDPH 6100?

Please describe the experience(s) briefly.

Who did you consult to come to your curriculum pathway
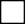
 1. M.D.-Ph.D. program director(s) decision? Please select all that apply.
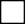
 2. Graduate school track leadership

3. Primary Ph.D. thesis mentor
4. Thesis advisory committee member(s)
5. Senior student(s)
6. Cohort classmate(s)
7. Residency program director(s)
8. Clinical mentor(s)
9. Dean(s)
10. Family / partner / friends
11. Other

If other, please describe.

My pathway informed or refined my clinical interests.

**Please rank how much you agree or disagree with each statement.**

**1 = strongly disagree, 2 = disagree, 3 = neutral, 4 = agree, 5 = strongly agree**

1. strongly disagree

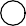

2. disagree	3. neutral	4. agree	5. strongly

agree

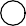

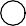

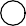

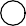

6. N/A

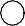

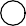

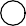

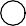

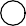

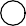

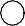
My pathway informed or refined my research interests.

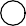

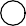

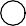

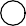

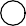

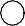
My clinical interests changed during my clerkship phase.

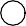

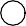

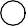

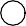

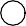

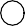
My clinical interests changed during my Ph.D. phase.

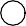

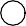

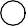

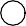

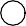

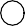
My research interests changed during my clerkship phase.

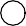
My research interests changed during my Ph.D. phase.

My clinical exposure informed my approach to research questions.

My research exposure informed my approach to clinical questions.

**Please rank how much you agree or disagree with each statement.**

**1 = strongly disagree, 2 = disagree, 3 = neutral, 4 = agree, 5 = strongly agree**

The length of my preclinical curriculum played a large role in my pathway decision.

1. strongly disagree

2. disagree	3. neutral	4. agree	5. strongly

agree

6. N/A

The intensity of my preclinical curriculum played a large role in my pathway decision.

Timing between Step 1 and Step 2 played a large role in my pathway decision.

Step 2 timing and scores played a large role in my pathway decision.

Clerkship grading played a large role in my pathway decision.

Letters of recommendation played a large role in my pathway decision.

Preparedness for post-clerkship rotations played a large role in my pathway decision.

Preparedness for away rotations played a large role in my pathway decision.

Continued training with my incoming cohort colleagues played a large role in my pathway decision.

The ability to participate in longitudinal clinical elective(s) played a large role in my pathway decision.

The ability to participate in a clinical re-entry program played a large role in my pathway decision.

My pathway allowed me to keep my clinical skills and/or knowledge fresh.

**Please rank how much you agree or disagree with each statement.**

**1 = strongly disagree, 2 = disagree, 3 = neutral, 4 = agree, 5 = strongly agree**

The timing of my required Ph.D. coursework / track requirements played a large role in my pathway decision.

1. strongly disagree

2. disagree	3. neutral	4. agree	5. strongly

agree

6. N/A

The requirements and/or timeline of my qualifying exams played a large role in my pathway decision.

The timing of my F30/F31 grant or other organizational/institutional grant played a large role in my pathway decision.

My desire to work on a particular project played a large role in my pathway decision.

The ability to remain engaged in laboratory work through my entire tenure in the program played a large role in my pathway decision.

Logistics related to publications played a large role in my pathway decision.

The ability to present at national/international meetings or conferences in my field played a large role in my pathway decision.

Having completed gap year(s) prior to matriculation played a large role in my pathway decision.

**Please rank how much you agree or disagree with each statement.**

**1 = strongly disagree, 2 = disagree, 3 = neutral, 4 = agree, 5 = strongly agree**

Having completed a masters degree prior to matriculation played a large role in my pathway decision.

1. strongly disagree

2. disagree	3. neutral	4. agree	5. strongly

agree

6. N/A

Personal reasons played a large role in my pathway decision.

The ability to pre-study for clerkships played a large role in my pathway decision.

The ability to TA for medical school course(s) played a large role in my pathway decision.

The ability to TA for quarter-long graduate school course(s) played a large role in my pathway decision.

The ability to TA for week(s)-long graduate or medical school course(s) played a large role in my pathway decision.

The ability to engage in leadership experience(s) played a large role in my pathway decision.

Is there a formal clerkship dedicated to your #1	1. Yes

**Please answer the following questions with your current top two desired residency / specialty**

**and future career plans. If you are undecided, please check undecided.**

choice desired residency / specialty?	2. No

3. Undecided

Is there a formal clerkship dedicated to your #2	1. Yes

choice desired residency / specialty?	2. No

3. Undecided

What are your plans for immediately after M.D.-Ph.D.	1. Residency

training?	2. Post-doctoral fellowship

3. Industry
4. Undecided
5. Other

Is your #1 choice desired residency / specialty	1. Yes

supported by at least one PSTP option?	2. No

3. Undecided

These include: Anesthesiology, Dermatology, Emergency Medicine, Internal Medicine, Pathology, Pediatrics, Psychiatry, Radiology, and Radiation Oncology.

Is your #2 choice desired residency / specialty	1. Yes

supported by at least one PSTP option?	2. No

3. Undecided

These include: Anesthesiology, Dermatology, Emergency Medicine, Internal Medicine, Pathology, Pediatrics, Psychiatry, Radiology, and Radiation Oncology.

Are you interested in pursuing a PSTP or similar	1. Yes

pathway?	2. No

3. Undecided

**Please rate your agreement or disagreement with the following.**

**1 = strongly disagree, 2 = disagree, 3 = neutral, 4 = agree, 5 = strongly agree**

My #1 choice desired residency / specialty is considered competitive.

1. strongly disagree

2. disagree	3. neutral	4. agree	5. strongly

agree

6. N/A

My #2 choice desired residency / specialty is considered competitive.

Competitiveness of my desired residency or specialty played a large role in my pathway decision.

Please share any additional thoughts / feedback / considerations related to the timing of your clerkships and Ph.D. phase.
