## Supplementary Appendix 2 for "Factors affecting trainees’ preferred timing of clinical clerkships during MD-PhD training"

Comparison of curricular pathway Likert scale-based ordinal data analyzed with chi-squared and Wilcoxon rank sum tests. Asterisks denote statistical significance at α=0.05 without correction for multiple comparisons. Results are separated by primary and secondary analyses, which stratified participants by traditional versus any clerkship prior to Ph.D. training (primary analysis) or 24- versus 18-month preclinical curriculum duration (secondary analysis).

| **ANALYSIS** | **DOMAIN** | **Item** | **χ^2^** | **p** | ***** | **rank sum** | **p** | ***** |
| --- | --- | --- | --- | --- | --- | --- | --- | --- |
| **Domain 1: Clinical training-related considerations** | | | | | | | | |
| Primary | 1 | 1 | 1.088 | 0.896 |  | 366.5 | 0.700 |  |
| Primary | 1 | 2 | 0.982 | 0.913 |  | 391.5 | 0.607 |  |
| Primary | 1 | 3 | 1.067 | 0.900 |  | 85.5 | 1.000 |  |
| Primary | 1 | 4 | 0.952 | 0.917 |  | 185.0 | 0.410 |  |
| Primary | 1 | 5 | 4.838 | 0.304 |  | 99.0 | 0.723 |  |
| Primary | 1 | 6 | 4.686 | 0.321 |  | 193.0 | 0.650 |  |
| Primary | 1 | 7 | 3.291 | 0.349 |  | 302.0 | 0.285 |  |
| Primary | 1 | 8 | 1.038 | 0.595 |  | 242.5 | 0.342 |  |
| **Domain 2: Graduate training-related considerations** | | | | | | | | |
| Primary | 2 | 1 | 4.808 | 0.308 |  | 410.0 | 0.124 |  |
| Primary | 2 | 2 | 3.483 | 0.481 |  | 314.5 | 0.676 |  |
| Primary | 2 | 3 | 1.644 | 0.801 |  | 331.5 | 0.488 |  |
| Primary | 2 | 4 | 1.849 | 0.764 |  | 313.5 | 0.322 |  |
| Primary | 2 | 5 | 3.846 | 0.427 |  | 274.5 | 0.377 |  |
| Primary | 2 | 6 | 5.407 | 0.248 |  | 271.5 | 0.745 |  |
| Primary | 2 | 7 | 5.255 | 0.262 |  | 378.5 | 1.000 |  |
| Primary | 2 | 8 | 6.280 | 0.179 |  | 393.0 | 0.124 |  |
| **Domain 3: Career development and personal considerations** | | | | | | | | |
| Primary | 3 | 1 | 3.850 | 0.427 |  | 450.0 | 0.152 |  |
| Primary | 3 | 2 | 6.701 | 0.153 |  | 434.5 | **0.047** | * |
| Primary | 3 | 3 | 6.522 | 0.163 |  | 383.5 | 0.235 |  |
| Primary | 3 | 4 | 5.253 | 0.262 |  | 370.5 | 0.927 |  |
| Primary | 3 | 5 | 3.294 | 0.510 |  | 402.5 | 0.892 |  |
| Primary | 3 | 6 | 3.988 | 0.408 |  | 290.5 | 0.229 |  |
| Primary | 3 | 7 | 3.289 | 0.511 |  | 453.5 | 0.119 |  |
| Primary | 3 | 8 | 0.682 | 0.953 |  | 319.5 | 0.910 |  |
| **Domain 4: Influence of pathway decision on framing interests** | | | | | | | | |
| Primary | 4 | 1 | 6.346 | 0.175 |  | 322.0 | 0.763 |  |
| Primary | 4 | 2 | 13.564 | **0.004** | * | 259.0 | 0.940 |  |
| Primary | 4 | 3 | 6.623 | 0.085 |  | 289.5 | 0.849 |  |
| Primary | 4 | 4 | 3.375 | 0.497 |  | 332.0 | 0.776 |  |
| Primary | 4 | 5 | 3.029 | 0.553 |  | 464.5 | 0.383 |  |
| Primary | 4 | 6 | 1.596 | 0.809 |  | 388.0 | 0.740 |  |
| Primary | 4 | 7 | 2.178 | 0.703 |  | 269.0 | 0.712 |  |
| Primary | 4 | 8 | 0.750 | 0.687 |  | 15.5 | 0.933 |  |

| **ANALYSIS** | | **DOMAIN** | **Item** | **χ^2^** | **p** | ***** | **rank sum** | **p** | ***** |
| --- | --- | --- | --- | --- | --- | --- | --- | --- | --- |
|  | **Domain 1: Clinical training-related considerations** | | | | | | | | |
|  | Secondary | 1 | 1 | 6.303 | 0.178 |  | 231.5 | 0.188 |  |
|  | Secondary | 1 | 2 | 3.195 | 0.526 |  | 302.5 | 0.840 |  |
|  | Secondary | 1 | 3 | 5.760 | 0.218 |  | 35.5 | 0.525 |  |
|  | Secondary | 1 | 4 | 9.716 | **0.045** | * | 186.0 | 0.925 |  |
|  | Secondary | 1 | 5 | 3.000 | 0.558 |  | 21.5 | 0.515 |  |
|  | Secondary | 1 | 6 | 3.705 | 0.447 |  | 181.5 | 0.755 |  |
|  | Secondary | 1 | 7 | 4.662 | 0.198 |  | 124.0 | 0.132 |  |
|  | Secondary | 1 | 8 | 1.100 | 0.577 |  | 171.5 | 0.512 |  |
|  | **Domain 2: Graduate training-related considerations** | | | | | | | | |
|  | Secondary | 2 | 1 | 2.556 | 0.635 |  | 238.0 | 0.814 |  |
|  | Secondary | 2 | 2 | 2.620 | 0.623 |  | 206.0 | 0.475 |  |
|  | Secondary | 2 | 3 | 5.294 | 0.258 |  | 203.5 | 0.290 |  |
|  | Secondary | 2 | 4 | 4.449 | 0.349 |  | 302.0 | 0.082 |  |
|  | Secondary | 2 | 5 | 7.523 | 0.111 |  | 315.0 | **0.007** | * |
|  | Secondary | 2 | 6 | 4.598 | 0.331 |  | 262.5 | 0.053 |  |
|  | Secondary | 2 | 7 | 6.530 | 0.163 |  | 227.0 | 0.146 |  |
|  | Secondary | 2 | 8 | 4.569 | 0.334 |  | 289.5 | 0.341 |  |
|  | **Domain 3: Career development and personal considerations** | | | | | | | | |
|  | Secondary | 3 | 1 | 1.873 | 0.759 |  | 287.5 | 0.791 |  |
|  | Secondary | 3 | 2 | 4.006 | 0.405 |  | 309.0 | 0.475 |  |
|  | Secondary | 3 | 3 | 2.491 | 0.646 |  | 316.0 | 0.208 |  |
|  | Secondary | 3 | 4 | 1.956 | 0.744 |  | 266.5 | 0.901 |  |
|  | Secondary | 3 | 5 | 2.438 | 0.656 |  | 290.5 | 0.867 |  |
|  | Secondary | 3 | 6 | 6.780 | 0.148 |  | 322.0 | **0.013** | * |
|  | Secondary | 3 | 7 | 2.395 | 0.664 |  | 308.5 | 0.690 |  |
|  | Secondary | 3 | 8 | 6.743 | 0.150 |  | 228.5 | 0.789 |  |
|  | **Domain 4: Influence of pathway decision on framing interests** | | | | | | | | |
|  | Secondary | 4 | 1 | 4.520 | 0.340 |  | 205.0 | 0.721 |  |
|  | Secondary | 4 | 2 | 0.144 | 0.986 |  | 190.5 | 0.943 |  |
|  | Secondary | 4 | 3 | 1.274 | 0.735 |  | 146.0 | 0.868 |  |
|  | Secondary | 4 | 4 | 2.178 | 0.703 |  | 205.0 | 0.557 |  |
|  | Secondary | 4 | 5 | 3.470 | 0.482 |  | 277.0 | 0.153 |  |
|  | Secondary | 4 | 6 | 5.345 | 0.254 |  | 235.5 | 0.074 |  |
|  | Secondary | 4 | 7 | 7.143 | 0.129 |  | 174.5 | 0.186 |  |
|  | Secondary | 4 | 8 | 3.333 | 0.189 |  | 7.0 | 0.300 |  |
