## Supplementary figures and images for "Factors affecting trainees’ preferred timing of clinical clerkships during MD-PhD training"

### Supplementary Figure 1

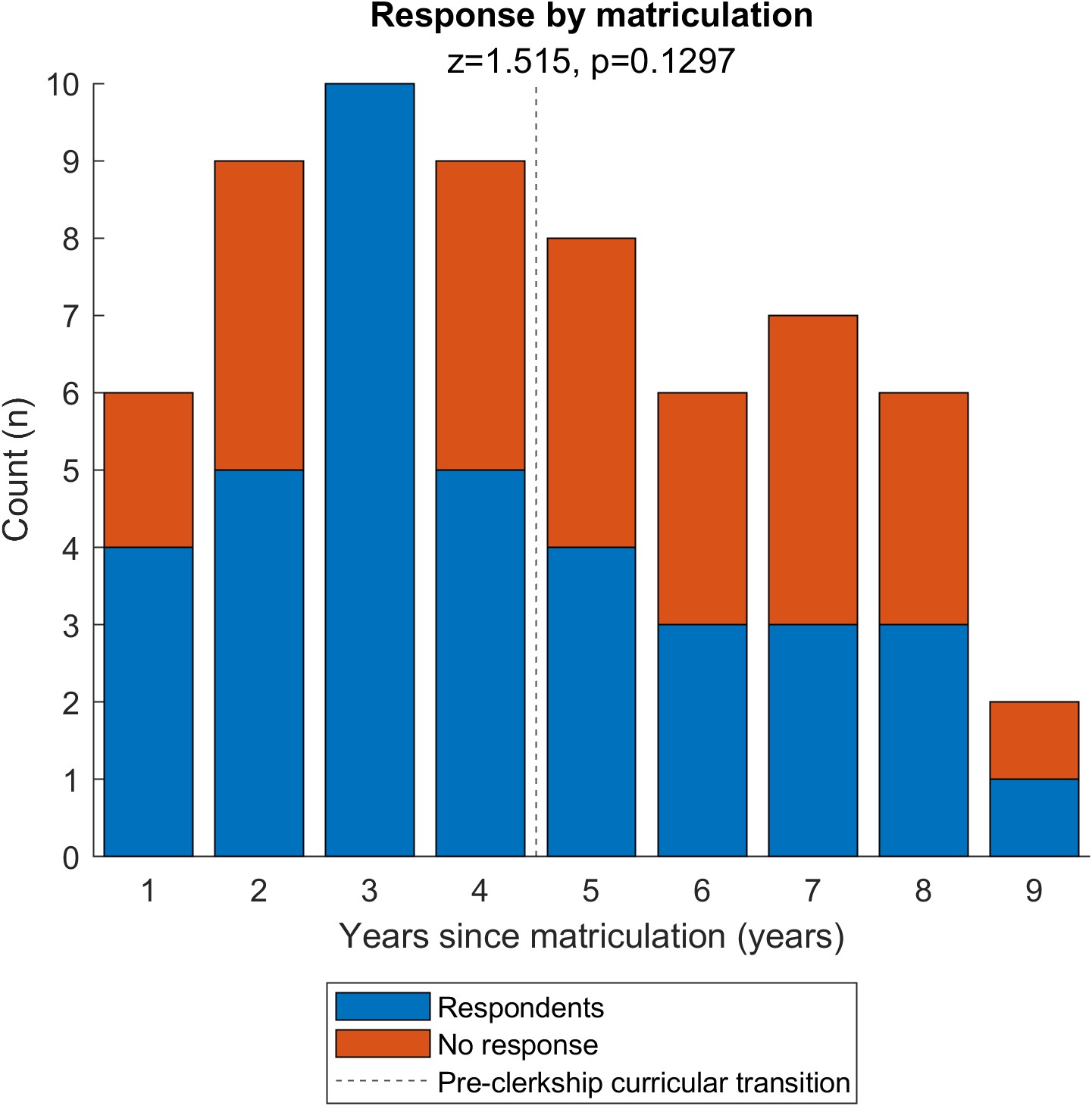
